# Exposomics and Cardiovascular Diseases: A Scoping Review of Machine Learning Approaches

**DOI:** 10.1101/2024.07.19.24310695

**Authors:** Katerina D. Argyri, Ioannis K. Gallos, Angelos Amditis, Dimitra D. Dionysiou

## Abstract

The principal objective served by this article is to identify key literature and provide an overview of the breadth of research in the field of machine learning applications on exposomics data with a focus on cardiovascular diseases. Secondarily, this study aimed at identifying common limitations and meaningful directives to be addressed in the future.

Most of the identified literature focuses on Disease Understanding, Prevention, and Management and the remaining on healthcare resource planning and management. Linear, non-linear, and ensemble machine learning methods have been applied, with tree-based methods generally being the most popular, and Random Forests specifically reported as being the best performer most of the time. Non-traditional CVD risk factors spanning behavioural/ lifestyle, socio-economic and a wide range of environmental factors have been investigated in the identified literature. According to our findings, the most reported category is the environmental risk factors which are often considered alone rather than in conjunction with the others.

Applications of machine Learning have been a substantial accelerating factor on the field, enabling the analysis of high dimensional data, improving accuracy, investigating novel risk factors and expanding our knowledge on the impact of exposome on cardiovascular diseases. However, several challenges persist, including data heterogeneity and the lack of standardized protocols, which introduces bias and hinders reproducibility and comparability across studies, as well as concerns about the validity of inference when applying machine learning methods to identify associations between exposure and health outcomes. Addressing these issues requires collaborative and interdisciplinary effort to integrate standardized exposome data frameworks akin to those of other –omics fields, apply causal inference methods to validate findings, and further expand the use of Explainable Artificial Intelligence to build insights and enable comparability and understanding.

## Introduction

Since the mid-20th century, cardiovascular (CV) diseases (CVDs) have emerged as the leading cause of death globally. In 2021, CVDs caused approximately 20.5 million deaths, accounting for approximately one third of all deaths worldwide [1]. In addition to this significant epidemiological burden, CVDs are estimated to impose a substantial financial burden, with the World Health Organization (WHO) recommending that health expenditures reach at least 5% of a country’s GDP [1]. They represent a large group of diseases attributed to a complex interplay between intrinsic risk factors, such as genetic predisposition, biological sex, and age, and lifetime exposure to environmental and behavioral risk factors which are considered at least partially modifiable [2]. Environmental exposures to ambient and indoor air pollution, noise, extreme temperatures, second-hand smoke, and chemicals, among other factors, have been recognized by many global health and environment agencies as significant contributors to the high burden of CVD. It is estimated that approximately 25% of deaths from Non-Communicable Diseases including primarily CVD-related deaths are attributable to environmental risks [2]. In recent years, there has been growing recognition of the importance of modifiable factors as a whole in efforts to alleviate the burden of disease [3].

While preventive interventions targeting traditional risk factors (e.g., blood pressure and cholesterol management) have aided in reducing CVD incidents, it remains a major global health issue, highlighting the need for new approaches. Unlike DNA, which remains unchangeable, there are primary modifiable contributors to CVD that are amenable to prevention and policy initiatives aimed at promoting cardiovascular health. The fact that environmental CV risk factors are inherently preventable leads to the actionable conclusion that reducing them is a key-step to alleviating the global burden of cardiovascular disease. In this context, investigations are increasingly directed towards non-traditional risk factors that are present in the built, natural, and sociο-economic environments comprising the “Exposome” [3], [4].

Exposome, the youngest member of the widely acknowledged -ome family, was first coined in 2005 by Dr. Christopher Wild, then-director of the International Agency for Research on Cancer (IARC), to complement the human genome and address the limitations of genetic research in explaining chronic disease etiology [5]. Aiming to fill this critical knowledge gap, the exposome was conceptualized as a systematic approach to measuring the entirety of environmental exposures encountered by an individual from conception onwards, including chemical, physical, biological, and lifestyle factors. In 2014, Gary Miller and Dean Jones expanded the exposome so as to emphasize diet, behavior, and endogenous processes, particularly focusing on biological responses to these exposures [6]. From their perspective, the exposome captures the essence of “nurture” in one of the oldest philosophical discussions of “nature” vs “nurture”, representing the summation and integration of external forces acting upon our genome throughout our lifespan. This includes factors such as diet, living environment, air quality, social interactions, lifestyle choices like smoking and exercise, and inherent metabolic and cellular activities. Measuring a quantity for the exposome serves as a biological index of our “nurture”, contextualizing the impact of specific exposures on health. This expansion and refinement of exposomics led to the inclusion of metabolomics moving beyond solely exposure-focused approaches to capture biological endpoints accompanied with substantial changes [7]. By exploring all these factors that constitute the exposome, researchers aim to understand and pinpoint modifiable risk factors and devise targeted interventions through active personal measures, behavioral strategies, novel policies, urban landscape reforms etc. to promote health and prevent disease across lifespan.

Along the lines of Genome-Wide Association studies (GWAS) and the identification of genetic basis of many complex traits and diseases [8], there have also been efforts to identify the “environmental” risk factors in the so-called ‘Environment-Wide association studies’ [9]. Finally, the wider term “Exposome-Wide Association Study” (ExWAS) has been proposed as a standardization term and a method designed to systematically investigate the connections between phenotypes and various exposures beyond the classic understanding of environmental factors, in line with the expanded definition of Exposome [6]. This approach facilitates the identification of significant correlations while addressing the challenge of multiple comparisons, aimed at finding the exposomic basis of a disease or trait [10]. Examples focused on CVD can be found in literature [11].

Different types of environmental risk factors have been reported in literature. First, chemical pollution, which spans air, soil, water and occupational pollution and is currently acknowledged as the most significant environmental cause of disease and premature death in the world [12]. Air pollution’s main health risks stem from particulate matter which is well known to be linked to CVDs [13], while the water pollution hazards arise from unsafe sources. The health impacts of soil pollution can be attributed mainly to heavy metals, deforestation, over-fertilization, and pesticides, with nano- and microplastics emerging as a threat. Although lead toxicity primarily results from water and soil pollution, it warrants separate consideration due to its widespread environmental presence. There is a close link between water and soil pollution, as polluted soil can contaminate surface and groundwater. Heavy metals and metalloids are particularly concerning due to their contribution to cardiovascular issues through oxidative stress and inflammation [14]. Chemical pollutants, particularly ambient air pollution, have garnered significant attention from research organizations assessing their impacts [15], [16]. These pollutants were also considered in the Global Burden of Disease study [17], [18].

Second, non-chemical pollution, such as transportation noise, light pollution and lack of green spaces, has been also shown to have substantial impact on CVD [4]. On the one hand, as urban areas expand and the demand for transportation increases, noise pollution is expected to rise. Research has demonstrated a connection between noise pollution and heightened CV risk, driven by mechanisms such as stress, sleep disruption, and increased inflammation. Furthermore, studies have shown an association between noise pollution and elevated risks of arterial hypertension, dyslipidemia, obesity, and type II diabetes mellitus. Thus, noise pollution not only directly impacts the CV system but also indirectly elevates the risk of developing traditional CV risk factors [19]. On the other end, it is known that nocturnal light pollution is associated with abnormal changes in circadian rhythms, which in turn may be linked to an increased risk of CVD [20], [21], increased blood pressure and risk of hospitalization for CVD [22]. While there is extensive and robust evidence linking noise pollution to an increased risk of CVD, the role of nocturnal light pollution in CV pathology is less studied. To validate and substantiate this association, further research is needed to figure out potential thresholds of ‘safe’ or ‘acceptable’ artificial light levels [23]. Finally, latest metanalyses have highlighted strong evidence on the link of green spaces and cardiovascular health [24]. For a comprehensive review on the epidemiology and pathophysiology of environmental stressors with a focus on CVDs, the interested reader may refer to pertinent literature [3], [4], [14], [25].

Apart from the classic environmental risk factors such as pollution, the exposome encompasses a wide range of lifestyle and socioeconomic factors. Growing evidence suggests that the CVD is characterized by socioeconomic inequalities [26], [27], [28]. These inequalities, most often assessed in terms of income, occupation and education, appear to be closely related with dietary habits and harmful lifestyle choices such as smoking and alcohol consumption [26]. Lifestyle interventions at targeting nutrition and physical activity have been shown to benefit cardiometabolic risk factors such as Body Mass Index (BMI), triglycerides and Low-Density Lipoproteins (LDL) in individuals at risk [29]. The overwhelming evidence of the impact of lifestyle on traditional cardiometabolic biomarkers has led to the emergence of the framework of “lifestyle medicine” which leverages lifestyle interventions to maintain cardiovascular health [30]. Most often these interventions focus primarily on diet, physical activity, perceived stress and anxiety and mitigation of harmful habits such as tobacco use and alcohol consumption.

While traditional epidemiology, which relies on well-established study designs, is a valuable tool for investigating the relationship between environmental exposures and health outcomes, the heterogeneous and dynamic nature of exposomics data presents new challenges. The exposome considers many environmental stressors simultaneously, as opposed to the one-by-one approach typically used in epidemiological research. This necessity, along with the large number of exposures, poses challenges such as increased complexity, high dimensionality, strong correlations between variables, and the need to understand both the combined effects and the causal structure between exposure risk factors and health outcomes. The Machine Learning (ML) toolbox, including Deep Learning (DL) techniques, is particularly well-suited to address these challenges [31]. By leveraging this set of tools, researchers can efficiently reduce data dimensionality and identify complex patterns and interactions; integrate diverse data types; enhance causal inference capabilities and uncover intricate relationships between exposomic factors and health outcomes; provide deeper insights into how combined environmental and behavioral factors influence CVD.

ML techniques, including DL, have been gaining popularity for quite a few years now in the analysis and integration of diverse kinds of -omics data (e.g., genomics, proteomics, metabolomics) especially within the context of precision medicine [32], [33]. However, applications on exposomics are still in early stages. This is partly because the field itself is relatively new (the term was only coined in 2005) but also due to the very nature and scale of the exposome which covers all exposures from conception to death. Thus, data is expected to exhibit significant heterogeneity (e.g., lifestyle factors, socioeconomic variables, biological responses etc.) as well as spatial and temporal variability, requiring integration from multiple sources and technologies. Due to these challenges, researchers proposed a roadmap for the use of federated approaches to accelerate research in the field [34]. Federated infrastructures and learning enable institutions to collaboratively analyze data while ensuring security and compliance with ethical, legal, and regulatory aspects. Leveraging federated technologies, researchers aim to utilize large, decentralized datasets and enable the sharing, access, and integration of diverse longitudinal exposomics data, supporting among others, ExWAS studies, functional exposomics, and precision health applications. Finally, DL, with its capacity for large-scale processing of complex and disparate multi-modal datasets, holds promise for advancing the understanding of the implications of the exposomics in the disease [35].

To date, to the best of our knowledge, the only review paper on the application of ML techniques on exposomics data for CVD-related investigations exclusively includes social determinants of health [36]. The primary objective of our work is to address this literature gap, by identifying key studies and providing an overview of the breadth of research in the field of ML applications on exposomics data with a focus on cardiovascular diseases. Additionally, we identify common limitations and propose meaningful directives for future research.

The remainder of the article is structured as follows: The Methods section reports the adopted literature search strategy and outlines the three research questions guiding our analysis. Subsequently, the Results section provides an overview of the relevant literature in the context of the three main axes of our work. Finally, we summarize our findings, discuss remarkable insights, and we identify and address the limitations of this study.

## Methods

### Search strategy

An extensive search of PubMed, IEEE Xplore and Google Scholar has been conducted to thoroughly identify articles on ML applications exploring potential associations of exposomics factors with CVD-related outcomes. In addition to the broad terms ‘exposome’ and ‘exposomics’, we derived key search terms based on the core aspects of the exposome as defined by the European Human Exposome Network [37]. The latter identifies environmental, lifestyle-related, and socio-economic factors as the key exposures constituting the exposome. These key factors have been adopted as the keywords for this review and searched within the titles and abstracts of published studies. Specifically, the body of work reported herein has been obtained using the following search query:

> *(“exposomics”[Title/Abstract] OR “exposome”[Title/Abstract] OR “environment”[Title/Abstract] OR “socio-economic”[Title/Abstract] OR “lifestyle”[Title/Abstract]) AND (cardiovascular disease[Title/Abstract]) AND (“machine learning”[Title/Abstract])*

Database searches have been supplemented with studies identified through manual searching. This review has been conducted using a systematic approach consistent with the Preferred Reporting Items for Systematic Reviews and Meta-Analyses (PRISMA) Extension for Scoping Reviews (PRISMA-ScR) [38]. No temporal bounds have been imposed on the publication dates.

### Study Exclusion Criteria

Overall, we have applied the following exclusion criteria to ensure that this review maintains focus and relevance to the desired scope:

- Non-English language articles,
- Non-peer reviewed items (e.g. gray literature such as pre-prints, technical reports, web-based guidelines etc.),
- Items for which the full text was not accessible (e.g., articles presented at conferences as abstracts),
- Articles exclusively focused on traditional statistical methods,
- Articles not exploiting exposomics data,
- Articles assessing intermediate risk factors such as blood pressure rather than direct CVD outcomes,
- Articles exploiting biomarkers as an input capturing endogenous metabolic changes that may result from various internal and external factors,
- Articles focusing on populations with pre-existing comorbidities (cancer, liver disease, etc.),
- Review articles,
- Duplicate articles.

### Research questions

The analysis to be presented revolves around the following Research Questions (RQ).

**RQ1**. How are studies categorized based on their primary use case, and which are their key distinguishing features?

**RQ2**. Which machine learning algorithms are more popular for investigating exposomics impact on CVDs and which seem to be ranking among the top performers?

**RQ3.** Which exposomic factors have been investigated and which have been identified as potentially good predictors?

The first research question (RQ1) focuses on categorizing the studies with respect to the context of application of the developed system. The second research question (RQ2) explores the ML algorithms preferred by the research community in an attempt to rank specific categories of algorithms with respect to their popularity in retrieved literature. Finally, the third research question (RQ3) aspires to shed light on specific categories of promising predictors and hopefully to identify new directions of exposomics variables investigations.

## Results

### A general overview

Firstly, an overview of the retrieved literature is provided, highlighting the temporal distribution of publications, the geographical distribution of utilized datasets, and the specific ML tasks addressed.

The timeline of publications identified by using the specified search criteria, as displayed in the bar plot below (Fig.1), reveals a pronounced increase of relevant studies from 2021 onwards.

**Figure 1.**
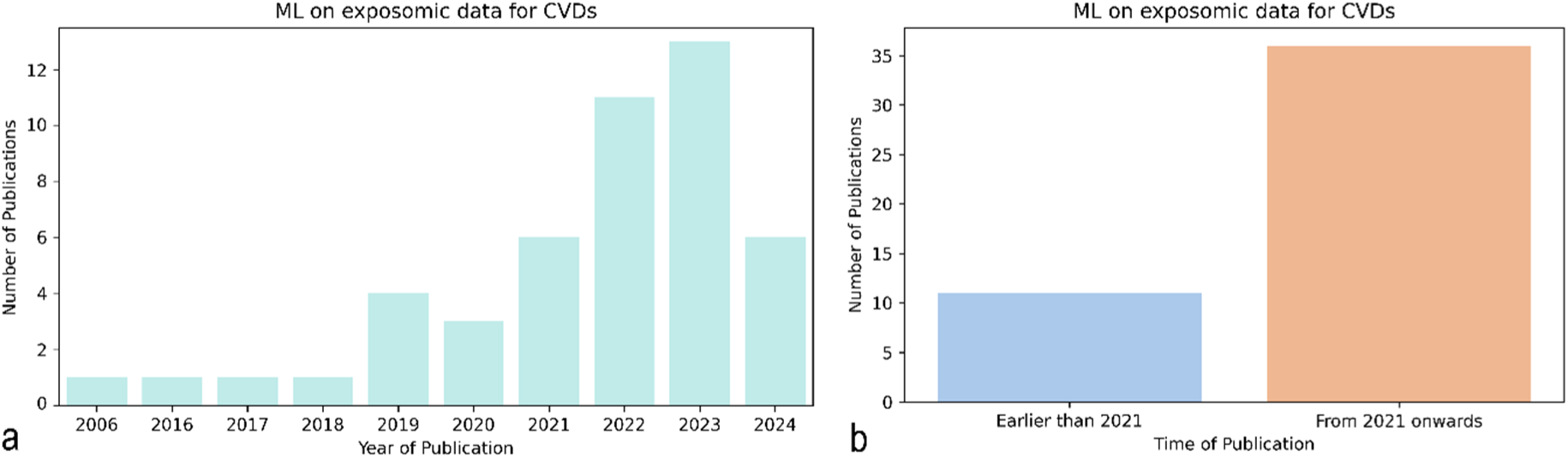
Time evolution of publications leveraging ML techniques to exploit exposomics datasets for CVD-related investigations a. Year of publication (x-axis) and number of identified studies per year (y-axis), b. Time of publication (‘earlier than 2021’ and ‘from 2021 onwards’)

Concerning the spatial distribution, the majority of identified studies reporting the origin of their datasets have used datasets collected within US or Asian territory (Fig. 2).

**Figure 2.**
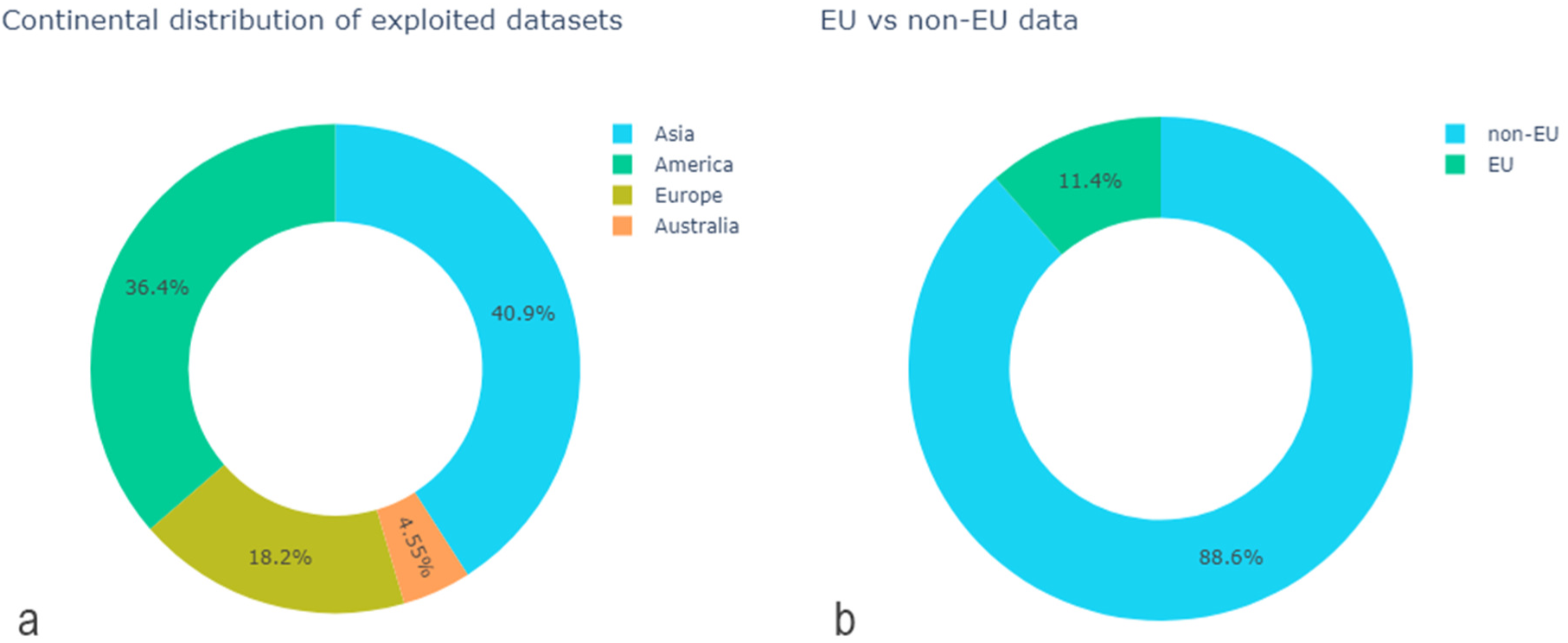
a. Continental distribution of exploited datasets, b. Data of EU vs non-EU origin.

Lastly, regarding the framing of the ML problems, an important observation is the pronounced, almost exclusive adoption of a supervised context. The most widely adopted problem framing is that of classification tasks, followed by regression tasks (Fig. 3).

**Figure 3.**
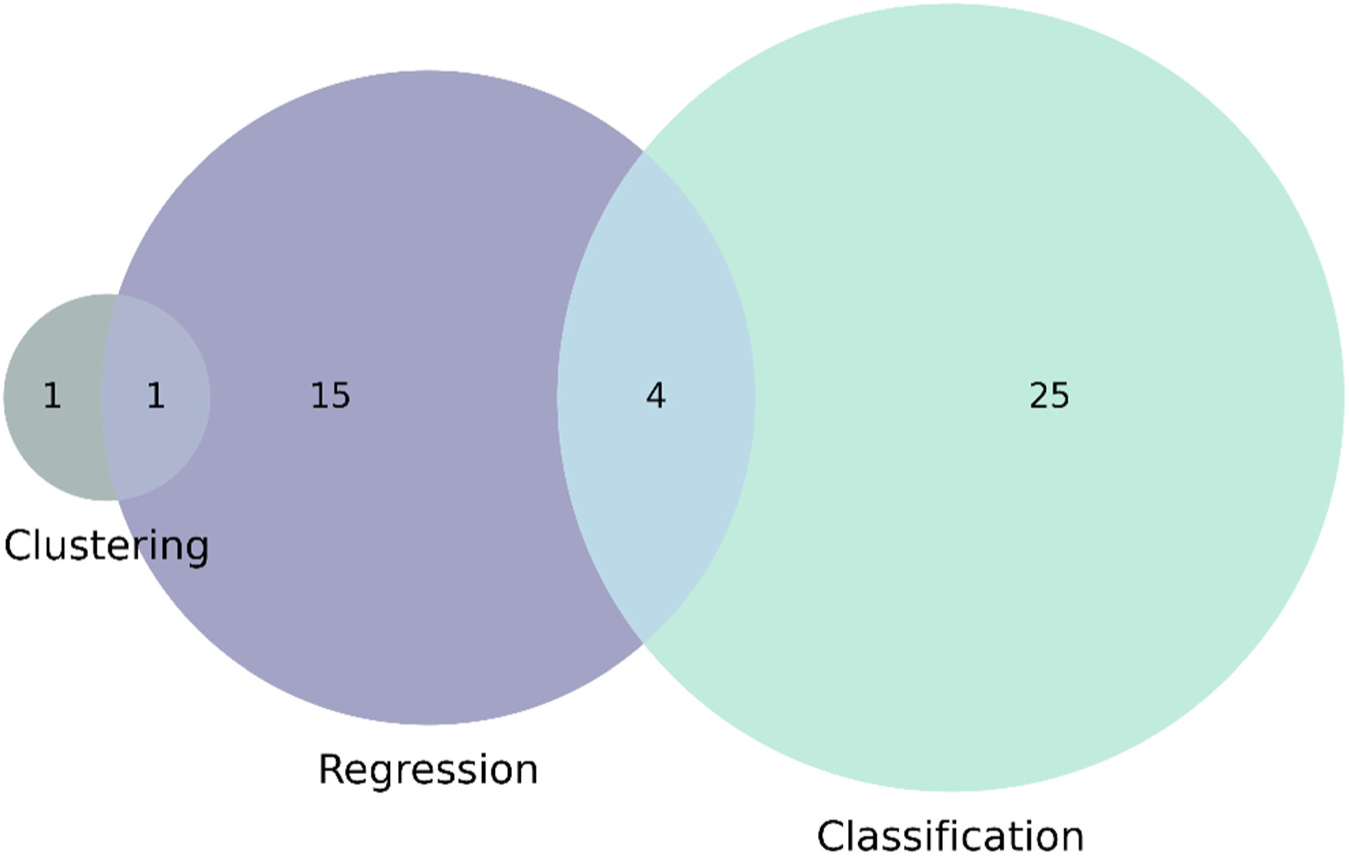
A Venn diagram of (core prediction) ML tasks categories encountered in literature focusing on exploring CVDs

### Research questions

Proceeding with the analysis conducted, we elaborate on identified categories of studies with respect to the aspects corresponding to the research questions:

RQ1. How are studies categorized based on their primary use case, and what are their key distinguishing features?

Two main categories of studies have been identified based on the application context. The first category focuses on disease understanding, prevention and management. These studies either pertain to direct citizen/patient care or inform population-level interventions and policy measures. The second category aims at assisting healthcare resource planning and management. It includes studies focused on forecasting hospital admissions, healthcare costs, and optimizing resources to ensure better system efficiency, and commonly aspires to build and eventually deploy an early-warning system for medical resource allocation and management.

Disease understanding, prevention and management are the primary focus of the retrieved studies. In this context, CVD incidence/prevalence or CVD-induced mortality are commonly used as target outcomes. The overall goal of these studies is to provide insights for reducing disease burden and mortality. A significant body of literature focuses on the development of personalized CVD risk assessment tools focusing on individual-level associations [39], [40], [41], [42], [43], [44], [45], [46], [47], [48], [49], [50], [51], [52]. In [53] this perspective is extended by exploring the link between maternal PM10 exposure during pregnancy and congenital heart defects in fetuses. Environmental Risk Score (ERS) measures (e.g. [54], [55]) have been proposed as a useful tool for characterizing cumulative risk from multiple exposures and are commonly used to estimate exposomic liability for a particular health outcome at an individual level. The multi-pollutant framework is gaining traction with the goal of better understanding the real-world health impacts of pollutant mixtures and facilitate risk stratification and targeted preventive interventions. It should be noted that several studies use the number of hospital admissions as a proxy to predict CVD incidence at a regional scale without considering aspects of healthcare system efficiency [56], [57], [58] and as such, we have considered them as part of the ‘Disease understanding, prevention and management’ category. Another portion of the literature deals with the prediction of CVD severity (e.g. [59], [60]) and CVD-induced mortality [61], [62], [63], [64]. As expected, significant focus is placed on the identification of novel risk factors for CVD incidence (e.g., [11], [65]) and CVD prevalence [35], [66] through the investigation of associations of variables of interest with selected outcomes. Another interesting research direction is the use of spatiotemporal data for spatially analyzing CVD prevalence [35], [66] or CVD-induced mortality (e.g. [60], [61]), and combining this knowledge with geographically targeted interventions, including urban-planning.

The second category, pertaining to healthcare resource planning and management, focuses on forecasting hospital admissions, healthcare costs, and optimizing resources to ensure better patient care and system efficiency. A key objective is the development of an early-warning system for healthcare resource allocation [67], [68], [69], [70]. Additionally, some studies address the financial sustainability of healthcare services, identifying key predictors of county-level healthcare costs [71] or high health-cost users among individuals with CVD [72].

**RQ2**. Which machine learning algorithms are found to be more popular for investigating exposomics impact on CVDs and which seem to be ranking among the top performers?

Tasks encountered in the retrieved literature have been predominantly addressed within classification and regression contexts, with clustering being much less common. Overall, a broad palette of ML algorithms spanning from linear to non-linear and ensemble algorithms has been exploited. In many cases different categories of algorithms are employed and compared against each other with respect to different evaluation metrics (e.g., accuracy in classification case, or MSE in regression). A detailed reporting of algorithms used in each study can be found in Table 1.

**Table 1.**
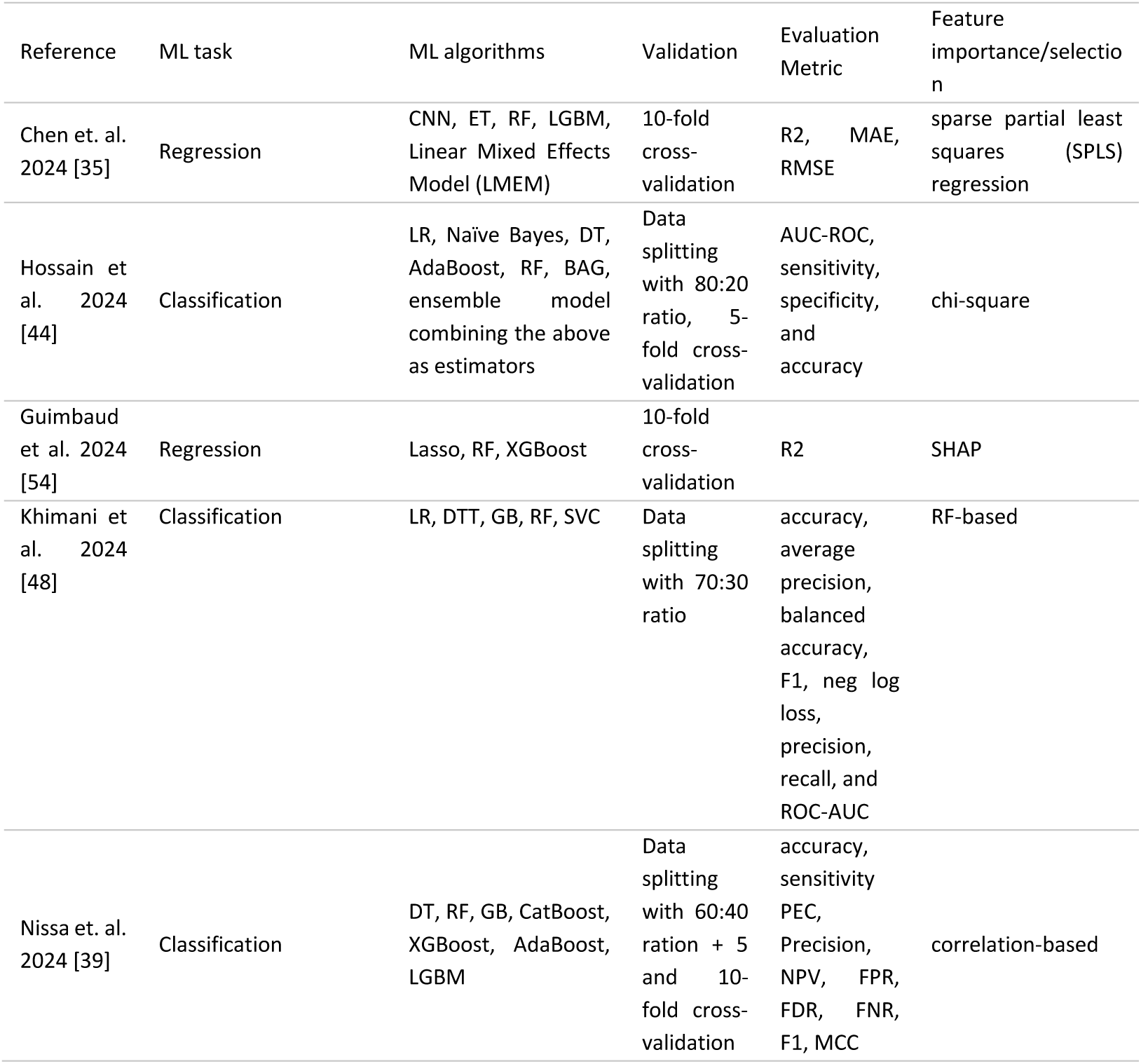
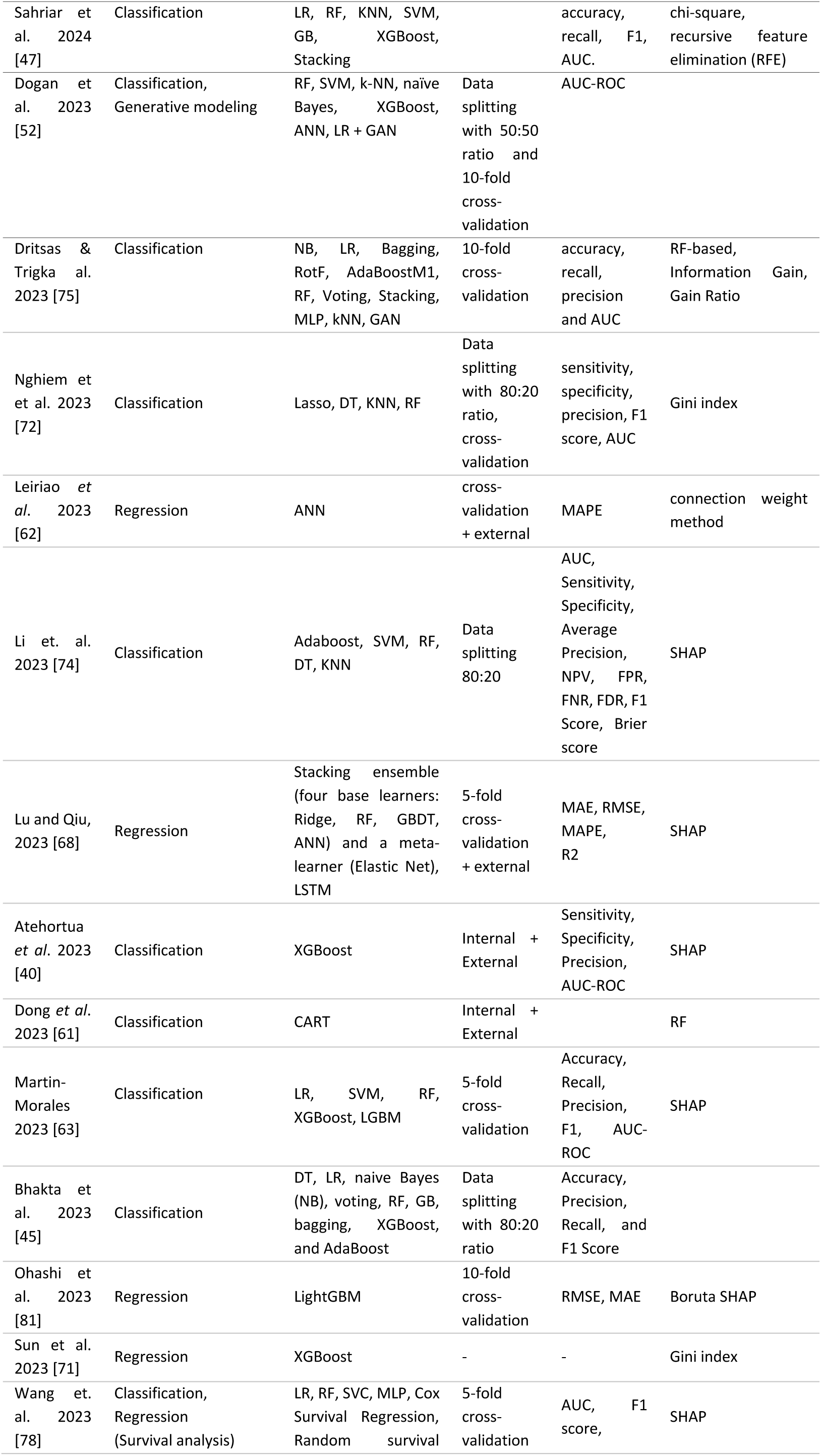
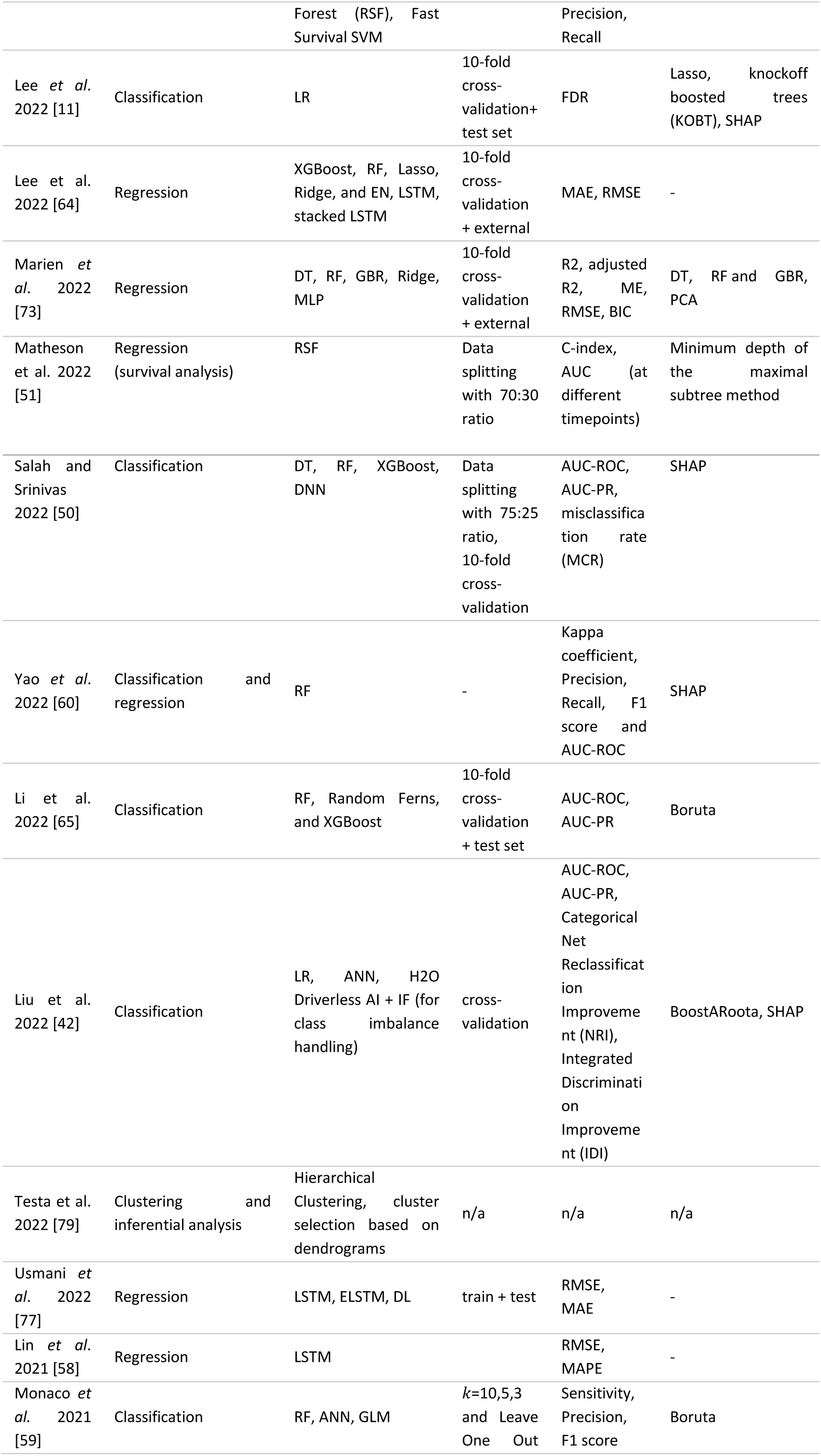
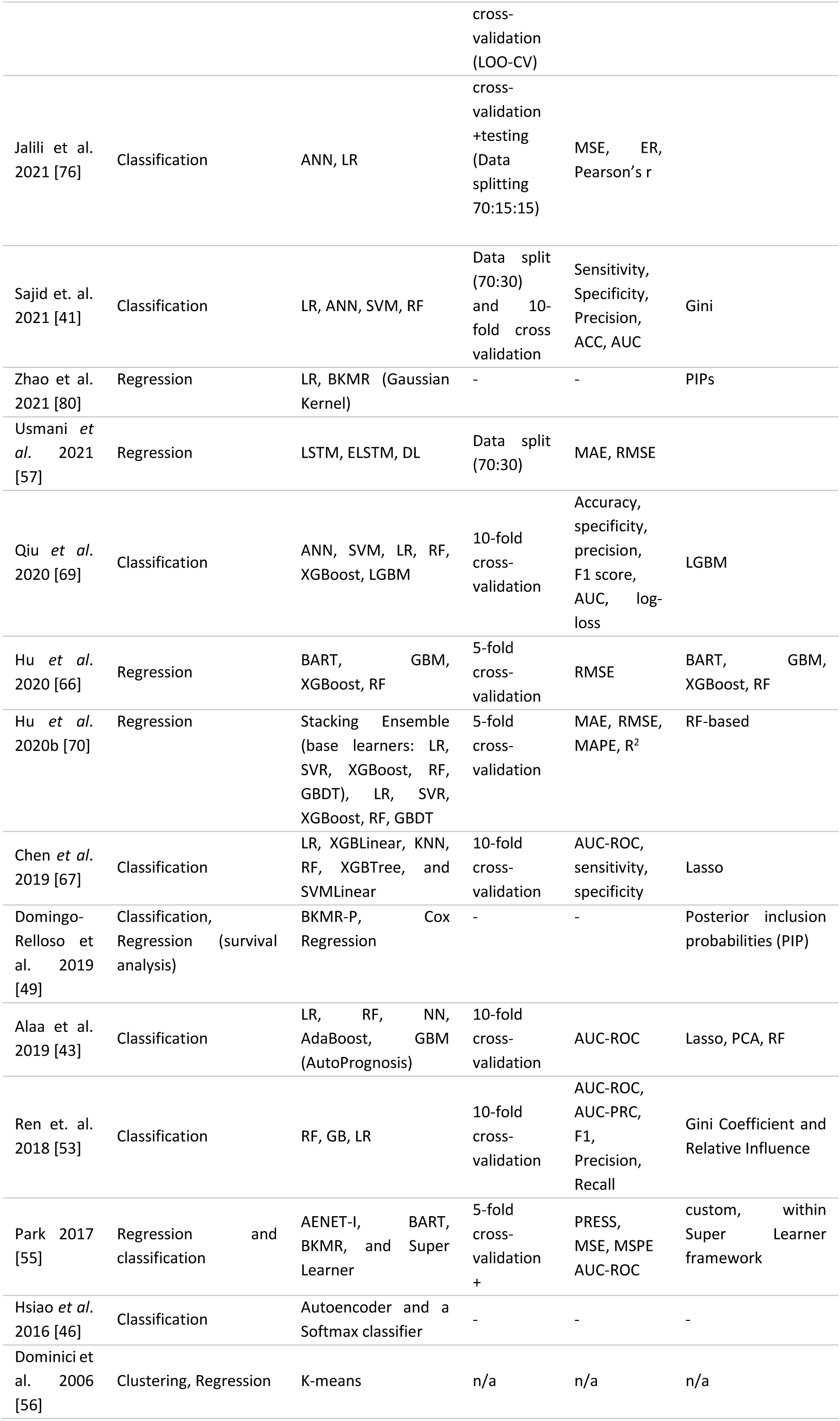
ML task addressed and key information on the adopted pipelines.

Research endeavors conducting comparative experiments among diverse algorithms account for nearly 70% of the identified studies. Starting from the most extensive investigations, a considerable part of the literature has explored a selection of linear, non-linear and ensemble methods [35], [41], [42], [44], [45], [47], [48], [52], [55], [64], [67], [68], [69], [70], [72], [73], [74], [75].

The second most commonly encountered comparative approach involves utilizing non-linear and ensemble methods [39], [50], [53], [54], [65], [74],have exploited linear and ensemble algorithms while [76] focused on linear and non-linear algorithms. Another comparative approach involves selecting more than one algorithm but within a specific family of models. In this context, [57] have focused on the use of non-linear models while a small portion of literature (e.g. [65], [66]) have exclusively investigated ensemble models.

Proceeding with the remaining 30% of the identified literature that does not conduct performance comparison whatsoever, a part of research opted for a non-linear algorithm [46], [56], [61], [62] while another part has focused on an ensemble algorithm of their selection [40], [51], [58], [60], [71].

As it becomes obvious from Figure 4 ensemble algorithms are the most popular choice [35], [39], [40], [41], [42], [43], [44], [45], [53], [54], [55], [59], [60], [63], [64], [65], [66], [67], [68], [69], [71], [72], [73] closely followed by non-linear algorithms [35], [39], [41], [42], [45], [53], [55], [55], [57], [58], [59], [61], [62], [63], [64], [65], [66], [69], [72], [73], [74], [76], [77], [78], [79]. The most widely adopted algorithms per category are Random Forest (RF) followed by Extreme Gradient Boosting (XGBoost) from the family of ensemble methods, Artificial Neural Networks (ANN) followed by Support Vector Machines (SVM) from the family of non-linear methods and linear/logistic regression as well as Lasso and Ridge variants from the linear algorithms.

**Figure 4.**
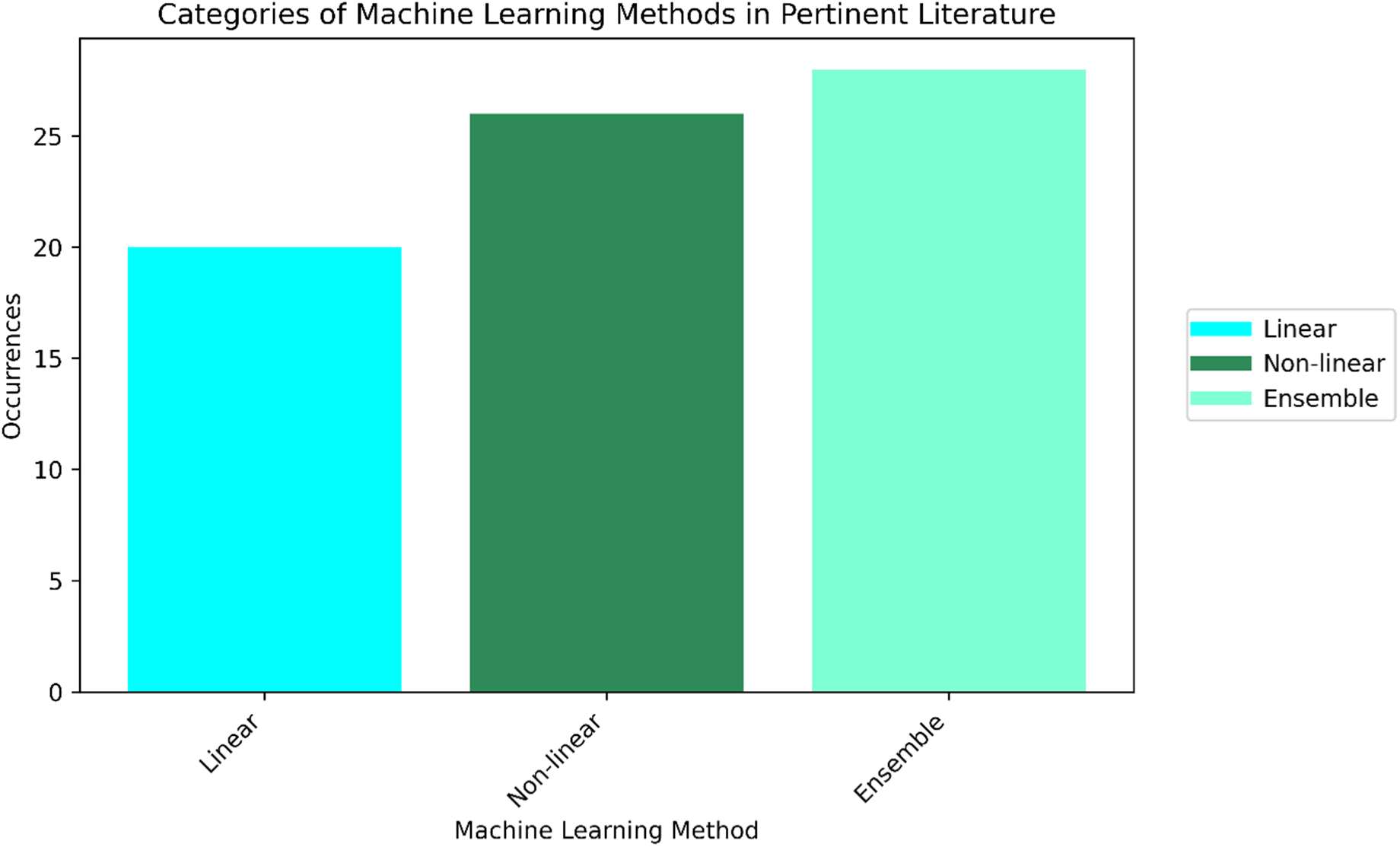
Popularity of categories of ML algorithms as reflected by the number of corresponding occurrences in identified literature.

As Table 2 shows, the majority of comparative studies identify different representatives of the family of ensemble models as top-performing algorithms. Among these, the RF algorithm is the most frequently highlighted as the best performer, followed by Stacking Ensemble.

**Table 2.**
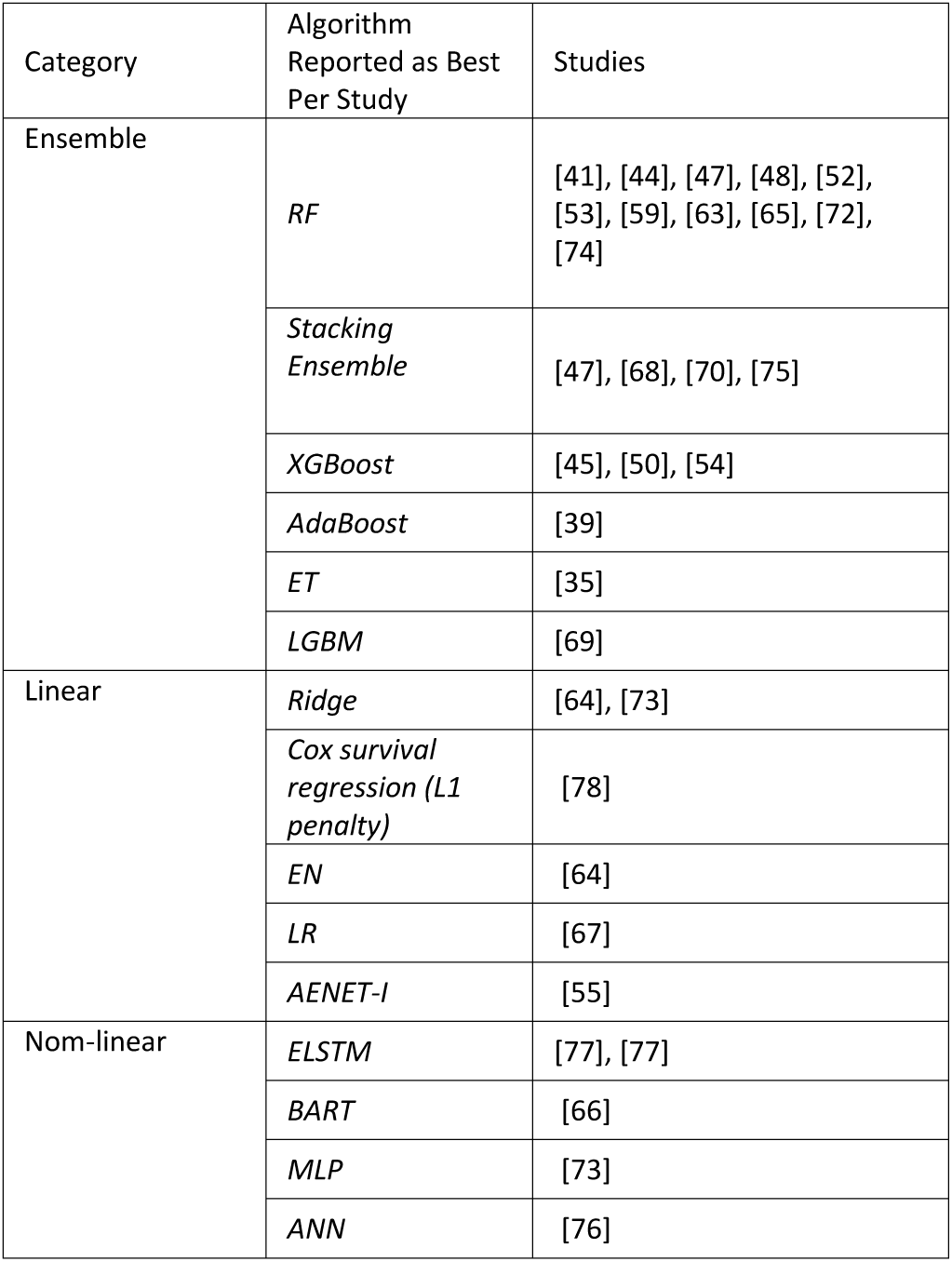
Best-performing algorithms per study.

| Category | Algorithm Reported as Best Per Study | Studies |
| --- | --- | --- |
| Ensemble | *RF* | [41], [44], [47], [48], [52], [53], [59], [63], [65], [72], [74] |
|  | *Stacking Ensemble* | [47], [68], [70], [75] |
|  | *XGBoost* | [45], [50], [54] |
|  | *AdaBoost* | [39] |
|  | *ET* | [35] |
|  | *LGBM* | [69] |
| Linear | *Ridge* | [64], [73] |
|  | *Cox survival regression (L1 penalty)* | [78] |
|  | *EN* | [64] |
|  | *LR* | [67] |
|  | *AENET-I* | [55] |
| Nom-linear | *ELSTM* | [77], [77] |
|  | *BART* | [66] |
|  | *MLP* | [73] |
|  | *ANN* | [76] |

Linear methods exploited in the relevant literature are linear/logistic regression [41], [42], [45], [48], [53], [63], [69], [76], [80], Linear Mixed Effects Models [35], SVM with linear kernel [67], Ridge regression [73], Lasso [54], [64], [72], Elastic Net (EN) [68], although the latter in a meta-learning context.

From the popular family of ensemble models, quite a few works have reported the use of Random Forests (RF) [51], [59], [60], [63], [65], [67], [69], [73], [74] with the latter using Random Survival Forest (RSF) i.e. an extension of the traditional Random Forest algorithm designed for survival analysis. Next widely adopted choices are XGBoost [40], [63], [65], [67] and Stacking ensemble model [68], [70], [75] followed by Light Gradient Boosting Machine (LGBM) [63], Gradient Boosting (GB) [73], Random Ferns [65], Bayesian Additive Regression Tree (BART) [55] .

Regarding non-linear methods, one of the most widely adopted methods is Support Vector Machines [41], [47], [48], [63], [69], [70], [74], [78]. Other algorithms commonly applied are kNN [47], [67], [72], [74], [75], Decision Trees [48], [50], Naive Bayes [44], [52] and Bayesian Kernel Machine Regression [49], [80]. Additionally, the use of artificial neural networks (ANN) and/or DL has been adopted in a considerable part of literature [35], [41], [42], [43], [46], [57], [59], [62], [69], [73], [76], [77]. It is noteworthy there is a single approach in the retrieved literature [52] where a GAN is also used model the joint distribution of risk factors, simulate possible disease progressions and to generate plausible alternative lifestyle scenarios and model uncertainties in disease progression.

**RQ3.** Which categories of exposomics factors have been investigated?

During the last years feature space investigations have broadened to include non-traditional CVD risk factors found in built, natural, and social environments, significantly contributing to the disease burden. The primary feature categories investigated in the retrieved literature, whether individually or in combination, are environmental exposure factors, lifestyle και socio-economic status-related factors. Most identified articles exploit exclusively environmental parameters while the remaining works consider either exclusively lifestyle factors or different combinations of these categories. For an overview see Fig.5 while for a more detailed reporting of the categories of exposomics features exploited in literature, see Table 3.

**Figure 5.**
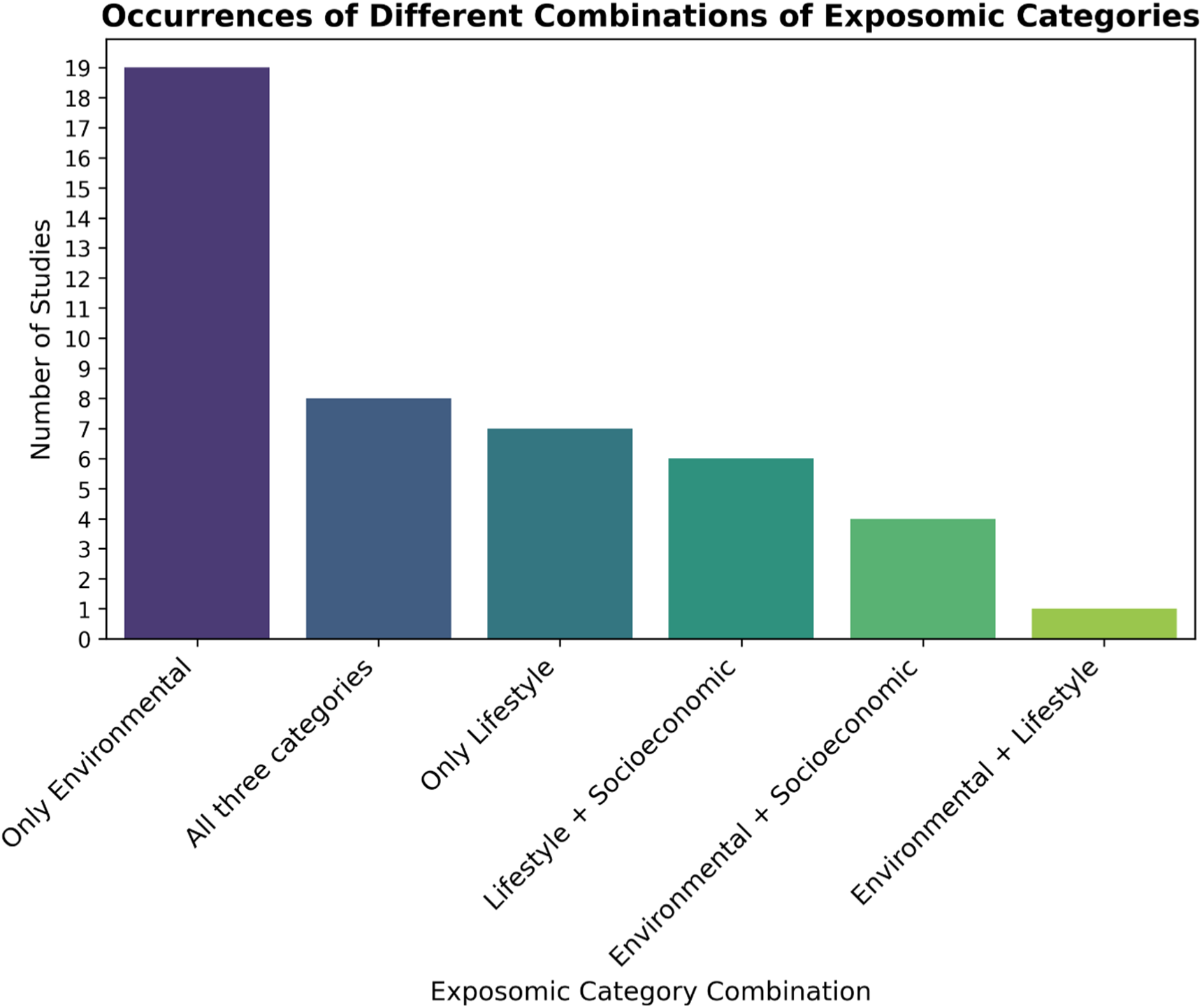
Occurrences of different combinations of exposomic categories used in retrieved literature.

**Table 3.** Exposome-related feature categories investigated and target outcomes addressed in pertinent literature.

| Reference | Exposomic categories included in feature space | Target outcome/variable |
| --- | --- | --- |
| Chen et. al. 2024 [35] | Socio-economic and, built environment | CHD prevalence |
| Hossain et al. 2024 [44] | Socio-economic, lifestyle | CVD incidence |
| Guimbaud et al. 2024 [54] | Air quality (pollutant concentrations), meteorological factors, traffic noise, | Cardiometabolic risk |
|  | traffic indicators, natural space, built environment, lifestyle, pollutant biomarkers |  |
| Kimani et al. 2024 [48] | Lifestyle | CVD incidence |
| Nissa et. al. 2024 [39] | Environmental, socio-economic and lifestyle factors (including diet, physical activity and sleep hours) | Risk of heart attack |
| Sahriar et al. 2024 [47] | Lifestyle, socio-economic, environmental | Stroke incidence |
| Bhakta et al. 2023 [45] | Occupation, environment, lifestyle habits | Heart disease detection |
| Dogan et al. 2023 [52] | Lifestyle | Coronary Heart Disease, MI, Stroke, any type |
| Dritsas & Trigka, 2023 [75] | Lifestyle | CVD incidence |
| Leiriao et al. 2023 [62] | Air quality, meteorological parameters | Cardiorespiratory mortality |
| Li et al. 2023 [74] | Exposure to heavy metals, socioeconomic | CHD Risk |
| Lu and Qiu, 2023 [68] | Air quality (pollutants concentrations) | Daily counts of hospital admissions (CVDs) |
| Atehortua et al. 2023 [40] | Early-life, environmental (noise/pollution levels), lifestyle, sociodemographics | Cardiometabolic risk estimation (coronary/ischaemic heart diseases, heart failure events, vascular dementia and cerebrovascular diseases) |
| Dong et al. 2023 [83] | Air quality (pollutants concentration), lifestyle, socioeconomic status | Premature CV mortality |
| Martin-Morales 2023 [63] | Dietary and non-diet-related health data | CVD mortality |
| Ohashi et. al. 2023 [81] | Daily weather data | CVD mortality |
| Sun et al. 2023 [71] | Demographic composition, risk factors, lifestyle and socioeconomic status | Total care costs per capita, inpatient care costs per capita, outpatient care costs per capita, CVD prevalence (%) |
| Wang et. al. 2023 [78] | Socioeconomic status, lifestyle (including dietary patterns) | CVD Mortality, IHD hospitalization |
| Salah and Srinivas 2022 [50] | Socioeconomic, lifestyle | CVD risk |
| Lee *et al.* 2022 [64] | Environmental (meteorological variables, air pollutants) | Non-accidental, cardiovascular, and respiratory mortality |
| Lee *et al.* 2022 [11] | Chemical and biological exposures, socioeconomic status, Lifestyle (food, diet and sleep) | Cardiac arrhythmia, congestive heart failure, coronary artery disease, heart attack, stroke, and a combined atherogenic-related outcome comprising angina, angioplasty, atherosclerosis, coronary artery disease, heart attack, and stroke incidents |
| Marien *et al.* 2022 [73] | Meteorological, air quality (pollutants concentration), vegetation index | Daily and annual CVD incidence (myocardial infarctions) |
| Matheson *et al.* 2022 [51] | Lifestyle | CVD incidence (CAD and ASCVD) |
| Yao *et al.* 2022 [60] | Urban multi-source spatio-temporal big data, road network, demographic, economic, meteorologic data and air pollutants | MI disease severity, MI mortality |
| Li *et al.* 2022 [65] | Socio-economic and lifestyle factors | CVD diagnoses |
| Liu *et al.* 2022 [42] | Demographic and lifestyle Information (Dietary Intake, Health Behaviors) | CVD incidence (Stroke) |
| Testa *et al.* 2022 [79] | Air Pollutants and weather variables | CVD incidence (Acute cardiac or cerebrovascular events) |
| Sajid *et. al.* 2021 [41] | Non-clinical factors such as education, area of living, occupation, exposure to economic problems and the role in the family | CVD risk |
| Usmani *et al.* 2022 [77] | Air quality (pollutants concentration) | Cardiorespiratory mortality |
| Lin *et al.* 2021 [58] | Air quality, meteorological | Emergency department visits (CVD-related etiology) |
| Monaco *et al.* 2021 [59] | Heavy metal concentrations (extracted by means of TMA hair tests.) | CVD risk |
| Jalili et al. 2021 [76] | Air quality (pollutants concentration), meteorological | CVD rate (as reflected by hospital admissions) |
| Usmani et al. 2021 [57] | Air quality (pollutants concentration) | Monthly cardiorespiratory hospitalization |
| Zhao et al. 2021 [80] | Dietary | ASCVD risk score (PCE) |
| Qiu et al. 2020 [69] | Air quality (pollutants concentration), meteorological | Peak demand days of CVDs admissions |
| Hu et al. 2020 [66] | Air pollution, lifestyle, dietary, socio-economic | Stroke incidents (neighborhood-level) |
| Hu et al. 2020 [70] | Air quality, environment | Daily number of Hospital Admissions for CVD |
| Chen et al. 2019 [67] | Air quality (pollutants concentration), meteorological | Hospital admissions (hemorrhagic stroke) |
| Alaa et al 2019 [84] | Health and medical history, lifestyle and environment, physical activity, psychosocial factors, dietary and nutritional information, and sociodemographics | CVD risk |
| Domingo-Relloso et al. 2019 [49] | Metal concentrations in urine | CVD incidence |
| Ren et. al. 2018 [53] | Air quality (pollutants concentration), meteorological (humidity, temperature) | Risk of Congenital Heart Defects |
| Park 2017 [55] | Metal biomarkers in blood and urine | CVD mortality |
| Hsiao et al. 2016 [46] | Air quality (pollutants concentration), meteorological | CVD risk |
| Dominici et al. 2006 [56] | Air quality (pollutants concentration), meteorological | Daily counts of county-wide hospital admissions (for primary diagnosis of cerebrovascular, peripheral, and ischemic heart diseases, heart rhythm, heart failure, chronic obstructive pulmonary disease, and respiratory infection, and injuries as a control outcome) |

Environmental exposure is the most widely adopted category of exposome [46], [49], [53], [55], [56], [57], [58], [59], [62], [64], [67], [68], [69], [70], [73], [76], [77], [79], [81] with the bulk of relevant literature exploring a broad palette of factors, ranging from pollutants concentrations (e.g. in [40], [46], [53], [54], [56], [57], [60], [61], [64], [67], [68], [69], [73], [76], [77], [79]), meteorological parameters ([46], [53], [56], [60], [62], [64], [67], [69], [73], [76], [79], [81]) to biomarkers reflecting the extent of human exposure to heavy metals ([49], [55], [59], [74]) concentration based on human scalp hair analysis, blood or urine samples. Less often, parameters reflecting noise pollution (e.g. in [40], [54]), green spaces ([54], [73]) and traffic proximity/volume (e.g. [40], [52], [66]) are also included in the exploited feature space. Lastly, a novel approach worth mentioning consists in the exploitation of machine vision-enabled assessment of the built environment to investigate potential associations with CVD-related variables [35]. Features extracted from Google Street View (GSV) images could also generate activation maps enabling the identification of high-risk neighborhoods.

Following closely are lifestyle-related factors, mostly reflecting dietary/sleep patterns along with various health habits. In this category, the focus is predominantly on health-related habits and adopted dietary (e.g. [11], [39], [40], [44], [50], [78], [80]), physical activity or the lack of it (e.g. in [43], [48], [50], [52]) and/or sleep patterns/quality (e.g. in [40], [50], [51], [52]). The most commonly considered health habit-related parameters is smoking status (e.g. in [47], [50], [52], [75]) and/or alcohol consumption (e.g. in [50], [52], [75]). In population-focused studies, group-specific input may also be incorporated. For example, in [50], a study focusing on adolescents, total screen time has also been included as a feature in the model.

Regarding socio-economic factors, a wide range of variables are used, primarily reflecting income, education levels, economic status and/or the subject’s occupation [11], [40], [42], [44], [45], [65], [72]. Less often, extra variables such as lack of private health insurance, home ownership, housing type or severe housing cost burden are also considered (e.g. in [61], [63], [66], [71], [78]).

A considerable part of literature explores combinations of all three categories i.e. environmental, lifestyle, and socio-economic factors [11], [39], [40], [43], [45], [61], [66]. Additionally, socio-economic and lifestyle-related parameters are frequently combined to investigate associations of exposomics data with CVD-related variables [44], [50], [65], [71], [78]. One last commonly encountered combination of categories consists in simultaneous investigation of socio-economic and environmental categories [35], [41], [60], [74]. Notably, socioeconomic are typically studied in combination with other categories.

## Discussion

This scoping review aimed at presenting the state-of-the-art of ML applications on exposomics data, specifically focusing on CVDs. To this end, a substantial amount of current literature on the selected topic has been identified and analyzed with respect to diverse aspects i.e. the use case of each application and their distinguishing features, ML techniques employed and the categories of exposomics data exploited. Building on the insights from the previous sections, we will summarize findings and elaborate on the strengths, the challenges and the opportunities that ML-enabled exploitation of exposomics data holds for CVD-related variables.

As reflected by the timeline of identified publications, the field is constantly expanding, particularly from 2021 onwards, with the expansion in exploitation of ML on exposomics targeting CVDs being predominantly driven by research in the US and Asia. However, the past 4 years the EU has shown an increasing interest in the Human Exposome Project initiating multimillion funding and thus recognizing potential to enhance public health [85].

Overall, ML approaches have provided novel insights into the exposomic factors affecting CVD by overcoming the inherent limitations of traditional epidemiological models, which assume linear and independent relationships between factors and outcomes. Unlike conventional regression-based models (such as linear regression and LASSO), which can only produce accurate interpretations under the assumption of linearity, ML techniques can capture nonlinear, synergistic, and threshold effects among complex exposures while also improving accuracy (e.g. in [44], [63]). This is particularly important in exposomics where high dimensional data is involved, and multiple factors interact in intricate ways that cannot be easily disentangled using standard statistical methods. By leveraging algorithms that are more robust to multicollinearity (e.g., random forests) and capable of identifying hidden patterns in high-dimensional data, ML enables the detection of previously unrecognized interactions [55].

As presented in detail in the context of RQ1, two main categories of studies have been identified based on the application context: the first one focusing on disease management, prevention and understanding, and the second one addressing healthcare resource planning and management. Overall, the direction of research efforts falls within the scope of AI-driven health investigations aimed at effective risk stratification, public health surveillance and health policy planning and the ML assisted exploratory association analysis between novel risk factors and CVD outcomes, all of which are described to contribute to health-related sustainable development goals [86].

Regarding ML techniques, it is noteworthy that ML tasks have been primarily framed in a supervised context, leaving ample space for the exploration of under-researched unsupervised techniques. Overall, a variety of ML algorithms spanning from linear to non-linear and ensemble algorithms have been exploited. In many cases different categories of algorithms are employed and compared against each other. Ensemble algorithms seem to be the most widely adopted approach and especially Random Forest, Stacking Ensemble and XGBoost which frequently rank as the most efficient solutions in comparative experiments. In the context of unsupervised learning and data clustering, more methods could be explored imposing minimal assumptions on data, trying to identify patterns, simplify large datasets and enhance predictive modeling. A valuable addition to the toolbox of the well-established linear clustering methods could be the case of non-linear clustering algorithms and specifically the so-called manifold learning, which aims to cluster data by identifying the intrinsic manifold upon which the data lies [87].

As presented in the context of RQ3, environmental exposure seems to be the most widely investigated aspect of the feature space followed by lifestyle-related exposure, with both of them often handled individually. In contrast, socioeconomic factors are typically studied in combination with other types of variables. Most environmental exposure studies are exploratory in nature, seeking to confirm associations of variables with health outcomes, instead of developing complex prediction models; they often aim to expand the understanding of the exposure-outcome relationships through identifying new or inadequately studied risk factors that may play an important role on disease prevalence and progression [11], [42], [65], [66]. Such efforts showcase the need for public health policies related to the environment [62].

A critical consideration in applying ML to exposomics research is the validity of inference. Many widely used ML algorithms, such as regression-based (e.g., LASSO) and tree-based (e.g., Random Forest and XGBoost) methods, are primarily designed for prediction rather than for modelling exposure-outcome associations (e.g., Bayesian Additive Regression Trees and Bayesian Kernel Machine Regression). Thus, they do not necessarily provide valid statistical inferences (e.g., they are often unable to quantify uncertainty around estimates) and/ or indicate true exposure-outcome associations. These algorithms are typically optimized with respect to prediction-oriented measures (e.g., AUC for classification, MAE for regression etc.) and then used to report associations - often relying on various feature importance metrics to identify key determinants [35], [39], [40], [41], [42], [44], [53], [55], [59], [60], [61], [62], [63], [65], [66], [68], [69], [71], [73], [74], [78], [81], [84], [88]. It is important to note that this approach raises concerns about post-selection bias, as feature selection is data-driven, and traditional statistical measures (e.g., p-values, confidence intervals) do not account for the fact that variables were selected based on observed data. For example, when conducting multiple statistical tests across many variables (e.g., in the context of linear regression tools), the likelihood of false positives increases, necessitating multiple testing adjustments (e.g., Bonferonni correction, False Discovery Rate etc.). Without proper correction, significance can be overstated resulting in potentially misleading conclusions. This is especially pertinent in exposomics, which frequently involve high-dimensional data with numerous, highly correlated exposures, and where identifying true predictors amidst correlated covariates is a significant challenge [89]. For example, in [62] the authors report the relative importance of each feature and compare the results with General Linear Model (GLM) where they consider the features significant based on p-value <0.05 without correcting for multiple comparisons.

Closely related to the above considerations is the issue of causal reasoning. When correlated variables are fed into ML models predicting CVD outcomes, their relative importance will be estimated higher when their impact on a predicted score is higher without necessarily implying causality. To enhance the reliability and generalizability of ML-driven exposomics research, a sound methodology should pay special attention to rigorous validation frameworks, including carefully chosen post-selection inference methods, such as cross-validation, permutation-based significance testing, stability selection, and, ultimately, external validation. However, the causality of the identified associations still needs to be validated independently within a causal inference framework [88] and/or through comparison with conventional association analysis [68]. When these aspects cannot be meticulously addressed, or are only partially addressed, authors should clearly acknowledge the limitations of their study. Expanding the use of causal inference methods (e.g., instrumental variable analysis, propensity score matching) within the ExWAS context, can help establish robust links between exposures and CVD outcomes, mitigating the impact of non-causal correlations and promoting the understanding of the underlying causal pathways.

Several additional limitations have been also reported across the identified literature. Firstly, the validity of the exploited ground truth cannot be ascertained, for example when using diagnostic codes (e.g., the international classification of diseases [65], [71]) corresponding to hospitalizations or death certificates [90]. Secondly, in the case of self-reported data categories such as lifestyle data [11], [39], [41], [42], [44], [45], [63], [78], [88], subjectivity and recall bias cannot be avoided. In addition, the bulk of conducted research only involved incidents that occurred in a single region and/ or the cohort was ethnically homogeneous [40], [44], [65], [71], [84]. Another important limitation stems from the lack of temporal alignment between the collection of medical data concerning CVD incidents and the collection of exposome-related data with many of the identified studies being observational in nature. Most of the time, the data sources were not comprehensive and/ or standardized [36]. Several works report limited availability of data sources [35], [44], [88], and finally, it is difficult to directly compare results from different studies since they are heterogeneous in terms of target, problem framing, study design, sample size and outcome assessment as reflected by the occasional lack of an external validation process but also by the diverse metric scores reported.

Most of these limitations are data-related, primarily arising from issues such as data heterogeneity, lack of standardization, inconsistent terminology, and incomplete, biased, or limited data sources. To address these challenges, it is essential to integrate standardized protocols for exposome data collection and harmonization across studies, reducing bias and enhancing reproducibility. Establishing a common language for exposomics is crucial for eliminating inconsistencies in terminology and classification. This would ultimately enable more effective comparison of findings across studies. It has been proposed by Schmitt et al. 2023 [34] that exposomics research would benefit significantly from a comprehensive, standardized, and interoperable data ecosystem framework, facilitating the integration of diverse longitudinal datasets across multidisciplinary [91] and international collaborations—similar to the Human Genome Project [92]. Such an ecosystem would link studies across continents, merging data from both existing and future longitudinal research. By establishing a federation of global data resources, this initiative would enhance the visibility, accessibility, and usability of exposomics data. Achieving this vision requires collaboration among multiple stakeholders, working together to form a global consortium that unifies frameworks and provides guidelines for an Exposome Study Design and a comprehensive exposome database [91]. In this context, it should be noted that the Environmental Exposure Assessment Research Infrastructure (EIRENE [93]) is the first European infrastructure dedicated to the human exposome.

The adoption of longitudinal study designs with synchronized environmental and health data collection can enhance the ability to track exposures over time and infer causality more reliably (e.g., the UK biobank utilized in [40], [84]). Additionally, integrating diverse data sources, including computerized and standardized data from wearables, can help mitigate recall bias, which is often introduced by self-reporting [11], [39], [40], [41], [42], [44], [45], [63], [65], [78], [84], [88]. Lastly, Explainable AI (XAI) techniques are valuable for enhancing model interpretability, enabling qualitative comparisons and ensuring that associations identified between exposomic factors and CVD-related outcomes are meaningful in real-world applications. It is noteworthy that many studies identified in this review implement XAI techniques in meaningful ways, attempting to interpret their models and explore potential associations (e.g., by using the model-agnostic SHapley Additive exPlanations) [11], [40], [42], [60], [63], [68], [74], [78], [81], [88].

All in all, the growing intersection of ML and CVD exposomics offers unprecedented opportunities to refine exposure assessment, improve early disease detection, and guide public health interventions. Expanding the feature space to include non-traditional CVD risk factors—spanning environmental, social, and lifestyle domains—has shown promise in enhancing the performance of conventional models targeting CVD-related variables. Such findings may serve as a foundation for developing high-precision tools for personalized healthcare management. On a broader scale, this approach holds potential for prioritizing healthcare resource allocation, optimizing workflows, and implementing targeted prevention and intervention strategies to reduce CVD-related healthcare costs.

However, to translate these advancements into actionable insights, stronger methodological rigor and standardization efforts are needed. Even though ML algorithms effectively enhance predictive modeling, most of them are not inherently designed for causal inference, raising concerns about the validity of reported exposure-outcome associations. Addressing limitations requires rigorous validation frameworks, careful post-selection inference methods, and integration of causal inference techniques into ML pipelines. Additionally, inconsistencies in data sources, lack of standardization, and the heterogeneous nature of studies hinder comparability and reproducibility. Establishing standardized protocols and interoperable data-sharing frameworks, akin to initiatives in other healthcare domains, holds the potential to significantly advance the field.

## Data Availability

This is a scoping review paper.

## Ethics declarations

### Human and Animal Rights and Informed Consent

This article does not contain any studies with human or animal subjects performed by any of the authors

## Abbreviations

AdaBoost: Adaptive Boosting
AENET-I: Adaptive Elastic-Net with main effects and pairwise interactions
AI: Artificial Intelligence
ANN: Artificial Neural Network
APS: Average Precision Score
ASCVD: Atherosclerotic Cardiovascular Disease
AUC-PR: Area Under the Precision Recall Curve
AUC-ROC: Area Under the Receiver Operating Characteristic Curve
AUC: Area Under the Curve
BAG: Bagging (regressor or classifier based on context)
BART: Bayesian additive regression tree
BKMR: Bayesian Kernel Machine Regression
BMI: Body Mass Index
CAD: Coronary Artery Disease
CART: Classification And Regression Tree CatBoost, Categorical Boosting
CHD: Coronary Heart Disease
CNN: Convolutional Neural Network
CVD: Cardio-Vascular Disease
GB: Gradient Boosting
DL: Deep Learning
DT: Decision Tree
ELSTM: Enhanced Long Short-Term Memory Model
EN: Elastic Net
ERS: Environmental Risk Score
ExWAS: Exposome-Wide Association Study
FA: Factor Analysis
FDR: False Discovery Rate
FNR: False Negative Rate
FPR: False Positive Rate
GAN: Generative Adversarial Network
GGT: Gamma-Glutamyl Transferase
GSV: Google Street View
IDI: Integrated Discrimination Improvement
IF: Isolation Forest
KNN: k-nearest neighbors
KOBT: Knockoff Boosted Trees
LASSO: Least Absolute Shrinkage and Selection Operator
LDL: Low-Density Lipoproteins
LGBM: Light Gradient Boosting Machine
LMEM: Linear Mixed Effects Model
LOO-CV: Leave-One-Out Cross-Validation
LR: Logistic Regression
LSTM: Long Short-Term Memory Model
MAE: Mean Absolute Error
MAPE: Mean Absolute Percentage Error
MCC: Matthew’s Correlation Coefficient
MI: Myocardial Infarction
ML: Machine Learning
MLP: Multi-Layer Perceptron
MSE: Mean-Squared Error
MSPE: Mean-Squared Prediction Error
ΝΒ: Naïve Bayes
NPV: Negative Predictive Value
NRI: Categorical Net Reclassification Improvement
PCA: Principal Component Analysis
PCE: Pooled Cohort Equations
PRESS: 
RF: Random Forest
RMSE: Root Mean Squared Error
RSF: Random Survival Forest
SHAP: SHapley Additive exPlanations
SVC: Support Vector Classification
SVM: Support Vector Machines
XAI: Explainable Artificial Intelligence
XGBoost: Extreme Gradient Boosting

